# Characteristics of COVID-19 in children and potential risk factors for requiring mechanical ventilation; an analysis of 22,490 cases from the United States

**DOI:** 10.1101/2023.02.06.23285543

**Authors:** Renuka Verma, Kamleshun Ramphul, Petras Lohana, Shaheen Sombans, Yogeshwaree Ramphul, Prince Kwabla Pekyi-Boateng

## Abstract

The pandemic of Coronavirus disease 2019 (COVID-19) has lasted more than two years and caused millions of deaths. While the characteristics and outcomes have been more widely studied in the adult population, we conducted an in-depth analysis via the 2020 National Inpatient Sample to understand the characteristics and predictors for the use of mechanical ventilation in patients of ages 18 and less in the United States. Twenty-two thousand four hundred ninety hospitalizations involving COVID-19-positive children were found. 52.7% (11850 cases) were females, 37.0% were Hispanics, 38.0% (8555 cases) were in the first percentile 0-25th of Median household income, and 66.9% used Medicaid. In total, 1140 cases (5.1%) needed mechanical ventilation. Among factors such as obesity (aOR 1.662, 95%CI 1.368-2.019, p<0.001), Blacks (vs. White) (aOR 1.472, 95%CI 1.23-1.761, p<0.001), private insurances (aOR 1.241, 95%CI 1.06-1.453, p=0.007) or remaining forms of payment other than Medicaid or private insurances (aOR 1.763, 95%CI 1.428-2.177, p<0.001, vs. Medicaid), ages 6 to 10 years (aOR 1.531, 95%CI 1.259-1.862, p<0.001, vs. ages 0-5) showed higher odds of needing mechanical ventilation. On the contrary, Females (aOR 0.54, 95%CI 0.472-0.617, p<0.001, vs. Males), hospitalized patients in November (aOR 0.542, 95%CI 0.399-0.736, p<0.001) and December (aOR 0.446, 95%CI 0.329-0.606, p<0.001) (vs. April), Hispanics (aOR 0.832, 95%CI 0.699-0.99, p=0.038, vs. White), ages 16-18 years (aOR 0.804, 95%CI 0.673-0.96, p=0.016, vs. 0-5years), and in the 76^th^-100^th^ median household income percentile (aOR 0.783, 95%CI 0.628-0.976, p=0.03, vs. 0-25th percentile) showed reduced odds. 9.6% of patients on mechanical ventilation died.

## Introduction

In late 2019, a new and atypical form of pneumonia was detected in China. After thorough analysis, a novel virus, Severe acute respiratory syndrome coronavirus 2 (SARS□CoV□2), was identified. The virus causes Coronavirus disease 2019 (COVID-19), which consists primarily of respiratory symptoms. It can, however, also influence other systems in the body.(1-4) As of December, there have been 653,219,490 cases of COVID-19 worldwide, costing the lives of 6,657,706 individuals (https://www.worldometers.info/coronavirus/).

Early reports associated a lower risk of mortality among children with COVID-19 compared to adults.(5) While the world eventually adapted over the last three years of the pandemic, there is a paucity of in-depth data and analysis among children. We, therefore, conducted a retrospective analysis to investigate further the outcomes and risk factors of COVID-19-positive children in the US that required hospitalization.

## Method

### Database

The National Inpatient Sample (NIS) is considered as one of the largest and most extensive all-payer inpatient database, covering over 35 million annual hospitalization records in the United States. The database contains cases of patients along with their diagnosis, procedures, and various clinically relevant characteristics in a de-identified model. It is produced each year by a partnership involving the Healthcare Cost Utilization Project (HCUP), Agency for Healthcare Research and Quality (AHRQ), and their collaborators across different states. More information and details related to access to the database can be found on https://www.hcup-us.ahrq.gov/nisoverview.jsp.(6)

### Analysis

We used the 2020 NIS for our study, which is the latest collection of NIS released to date. Since the ICD-10 diagnosis code for COVID-19 (U071) was released on April 1st and has high sensitivity and specificity, we included only cases between April 1st, 2020 to December 31st, 2020 in our study.(7) Children aged 0 to 18(inclusive) were recruited. Several patient characteristics, such as age, sex, race, region, and month were compared using Chi-Square tests. Appropriate ICD-10 codes from HCUP allowed us to include several potential comorbidities. The presence of mechanical ventilation was also studied, and logistic models adjusting for diverse patient variables allowed us to estimate their adjusted odds ratio (aOR). Mortality rates were also calculated. SPSS 29 (Armonk, NY: IBM Corp) was used to conduct all our analyses. Finally, as HCUP recommends using the discharge weight (DISCWT) to extract a national estimate, we report all our results as adjusted in weighted form for improved accuracy.(6)

### Ethic clearance

Since the patient information from NIS is in de-identified form, HCUP waives users from requiring IRB and other forms of ethical clearance for its use. However, users who had access to the database were mandated by HCUP to undergo their training, and to sign the appropriate DUA from the providers.(6)

## Results

### Basic Patient characteristics

Our study found 22490 cases of COVID among children between the ages of 18 and less in the United States that required hospitalized care between April 1st, 2020 to December 31st, 2020. 52.7% (11850 cases) were females, while 47.3% (10640 cases) were males. 34.2% were between the ages of 0-5years, 11.0% were between 6 to 10 years, 22.4% were between 11 to 15 years, and 32.4% were aged 16 to 18 years. The mean age was 9.77 years. Racially, 27.6% were Whites, 19.9% were Black, 37.0% were Hispanics, and 9.1% were races other than White, Black or Hispanic. The most common insurance provider was Medicaid, as 66.9% of cases were covered. Moreover, 25.7% of cases were covered via private insurance forms, and other forms of payments covered 7.4%.

Furthermore, 38.0% (8555 cases) were in the first percentile 0-25th of Median household income, with 25.5% (5740 cases) among the 26th-50th percentile, 20.8% (4685 cases) in the 51st to 75th percentile, and 14.3% (3220 cases) were classified as 76th to 100th percentile. Several comorbidities and risk factors were also seen. 5.9% of cases had depression, 10.7% were obese, and 13.0% had asthma. One thousand one hundred forty cases (5.1%) required mechanical ventilation during their hospitalization with COVID. The mean length of stay was 5.28 days, the mean hospital charge was $70650.29, and the total charge was estimated at $1,557,838,670. Unfortunately, 140 deaths (0.6%) were also recorded in patients of ages 18 or less admitted with COVID.

### Mechanical ventilation

Various predictors and risk factors that could lead to the need for mechanical ventilation among COVID-19-positive children were identified in our analysis (table 1). Females were less likely than males to require mechanical ventilation (aOR 0.54, 95%CI 0.472-0.617, p<0.001), while obesity showed increased odds (aOR 1.662, 95%CI 1.368-2.019, p<0.001). As COVID-19 progressed over 2020 in the US with multiple waves, patients admitted in November (aOR 0.542, 95%CI 0.399-0.736, p<0.001) and December (aOR 0.446, 95%CI 0.329-0.606, p<0.001) showed lower odds of requiring mechanical ventilation than those admitted in April. Racial differences were also observed as those classified as Blacks (aOR 1.472, 95%CI 1.23-1.761, p<0.001) were more at risk than White, while Hispanics (aOR 0.832, 95%CI 0.699-0.99, p=0.038) showed lower risks. Furthermore, those covered by private insurance (aOR 1.241, 95%CI 1.06-1.453, p=0.007) or any other forms of payment (aOR 1.763, 95%CI 1.428-2.177, p<0.001) other than Medicaid or private insurances, had increased odds of mechanical ventilation (vs. Medicaid).

**Table 1.** Adjusted odds ratio (aOR) of requiring mechanical ventilation among patients ages 0-18 admitted with COVID-19 in the United States in 2020.

| Characteristics | 95% Confidence Interval |  | aOR | P-value |
| --- | --- | --- | --- | --- |
|  | Lower | Upper |  |  |
| Month |  |  |  |  |
| April | Reference |  | 1 |  |
| May | 0.595 | 1.146 | 0.826 | 0.252 |
| June | 0.791 | 1.479 | 1.081 | 0.625 |
| July | 0.645 | 1.168 | 0.868 | 0.35 |
| August | 0.779 | 1.423 | 1.053 | 0.735 |
| September | 0.626 | 1.205 | 0.868 | 0.398 |
| October | 0.664 | 1.254 | 0.912 | 0.572 |
| November | 0.399 | 0.736 | 0.542 | <.001 |
| December | 0.329 | 0.606 | 0.446 | <.001 |

| Patient characteristics and risk factors |  |  |  |  |
| --- | --- | --- | --- | --- |
| Female | 0.472 | 0.617 | 0.54 | <.001 |
| Obesity | 1.368 | 2.019 | 1.662 | <.001 |
| Depression | 0.998 | 1.749 | 1.321 | 0.051 |
| Asthma | 0.914 | 1.318 | 1.097 | 0.321 |
| Race |  |  |  |  |
| White | Reference |  | 1 | <.001 |
| Black | 1.23 | 1.761 | 1.472 | <.001 |
| Hispanic | 0.699 | 0.99 | 0.832 | 0.038 |
| Other Races | 0.732 | 1.205 | 0.94 | 0.624 |
| Primary payer form (insurance form or payment method) |  |  |  |  |
| Medicaid | Reference |  | 1 |  |
| Private Insurance | 1.06 | 1.453 | 1.241 | 0.007 |
| Other forms of payment | 1.428 | 2.177 | 1.763 | <.001 |
| Age group |  |  |  |  |
| Age 0-5 years | Reference |  | 1 |  |
| Age 6-10 years | 1.259 | 1.862 | 1.531 | <.001 |
| Age 11-15 years | 0.747 | 1.083 | 0.899 | 0.262 |
| Age 16-18 years | 0.673 | 0.96 | 0.804 | 0.016 |
| Median Household income quartile |  |  |  |  |
| 0-25th | Reference |  | 1 |  |
| 26th-50th | 0.862 | 1.195 | 1.015 | 0.861 |
| 51st- 75th | 0.931 | 1.31 | 1.105 | 0.254 |
| 76th to 100th | 0.628 | 0.976 | 0.783 | 0.03 |

Children between the ages of 6-10 years (aOR 1.531, 95%CI 1.259-1.862, p<0.001) had higher odds of needing mechanical ventilation than those of ages 0-5, while those between 16-18 years (aOR 0.804, 95%CI 0.673-0.96, p=0.016) showed reduced odds (vs. 0-5years). Finally, socioeconomic differences were also noted as those in the 76th to 100th percentile (aOR 0.783, 95%CI 0.628-0.976, p=0.03) were less likely to require mechanical ventilation than those from the 0-25th percentile of income. The mean age of patients requiring mechanical ventilation was 9.94 years (vs. 9.76, p<0.01). 110 (9.6%) patients that required mechanical ventilation died (vs. 0.1% that did not require it).

## Discussion

Our study from the 2020 NIS provides a new perspective on various data for children admitted for COVID-19 during the first year of the pandemic in the United States. A higher percentage of patients were females compared to males in our study, which is different from the previous study reported with a smaller sample size by Parcha et al..(8) Furthermore, a higher proportion of the COVID-19-positive children were racially classified as Hispanic. This correlates with previous reports that have strongly hinted towards strong racial and ethnic differences in risk and even outcomes.(9) The socioeconomic and higher prevalence among the lower income quartile highlights the socioeconomic differences proposed previously by Goyal et al., who also hypothesized that living in multigenerational households can also increase their exposure via older adults who are positive.(10) Finally, the mortality rate among patients ages ≤18 years (0.6%) was lower than the rate among patients ages ≥18 years reported by Isath et al. (13.2%).(11)

In the second part of our analysis, we found that patients requiring mechanical ventilation had higher odds of mortality and that many potential factors influenced their odds of requiring mechanical ventilation. Several early publications categorized patients as requiring mechanical ventilation in the severe group of COVID-19, and they have all shown a poor outcome.(12-14)

Higher odds of mechanical ventilation were seen in April compared to November and December. This can correlate with the start of the pandemic in the US and the adaption phase and changes in protocols that had to be tried and tested during that time. Furthermore, it may reflect the hesitancy to seek care in the early phase of the pandemic, which may be due to any social stigma, fear, or false belief due to misinformation by the parents.(15-18) Further study should be carried out to properly understand the differences, as this can help in future pandemics. Moreover, differences between the variants in April and late 2020 and their impact on children should also be compared.(19)

Females in that age group that were COVID-19-positive also had lower odds of needing mechanical ventilation. Various factors can influence the outcome based on differences in sex, as males and females have exposure to different social and health risk factors.(20) Our analysis also found that patients with obesity had higher odds of needing mechanical ventilation. Similar results have been found among the adult population.(21, 22) Obesity can trigger various immunological pathways via the varying levels of cytokines that predispose individuals to a poorer outcome when admitted with COVID-19.(21, 22)

Differences based on age groups were also seen as children of ages 0-5 had higher odds of needing mechanical ventilation than those of 16-18 years, but lower compared to those of ages 6-10 years. In their meta-analysis, Harwood et al. reported a higher risk of critical care admission in younger children.(23) Similar results were described by Preston et al., who found that children ages 2-5 and 6-11 had higher odds of severe COVID-19 than those ages 12-18.(24)

Finally, our analysis also highlighted several racial and socioeconomic disparities that influenced the requirement for mechanical ventilation. While a higher proportion of all COVID-19 cases in our study were Hispanics, they showed lower odds than Whites of requiring mechanical ventilation. On the other hand, Blacks had higher odds. Such findings confirmed previous reports of similar racial disparities.(24, 25) Medicaid insurers had lower odds of requiring mechanical ventilation than those covered by private insurances, or other forms of insurances. A recent report by Rakus et al. described the importance of Medicaid in covering various low-income populations that may have contributed to lowering the impact of the pandemic.(26) However, further analysis should be encouraged to understand the impact on children, as seen in our study. Another significant socioeconomic disparity that we found consisted of the higher odds of mechanical ventilation among those who are from the lower quartile of median household income compared to those from the top quartile. Access to care during the pandemic, as well as timely education on the importance and early identification of the symptoms, may have impacted these groups. Similar disparities have been reported worldwide.(27-29) Global initiatives should focus on lowering these gaps to lower the impact based on economic backgrounds.

While our study was able to report several details on the hospitalization of patients of ages ≤18 years for COVID-19 in the United States, there were some limitations to it. We could not report these patients’ long-term progress and monitor their condition after they have been discharged. Furthermore, coding mistakes in hospital settings are common, and may influence the values presented. However, our study provides one of the most extensive analyses for that age group. We encourage future researchers to explore and understand the various results and compare them to their clinical settings. The pandemic of COVID-19 has shown the world that we are vulnerable to a rapid fall and brought much stress to the healthcare system. Therefore, it is vital to use the pandemic as an example to improve access to education and treatment for future outbreaks or pandemics globally.

## Conclusion

Between April 2020-December 2020, 22490 hospitalizations among children ages ≤ 18 years in the United States were reported. Children in our study reported a lower mortality rate than adults, as per past studies. We also identified several racial and socioeconomic differences among the patients. Furthermore, differences in factors influencing the odds of needing mechanical ventilation were also seen. Patients using mechanical ventilation had a poorer prognosis.

## Data Availability

The data used in this study can be found on the HCUP website. Anyone interested with the use of the data can request access to HCUP directly via their respective DUA and rules as per https://www.hcup-us.ahrq.gov/db/nation/nis/nisdde.jsp. The authors are not allowed to share the data directly.

https://www.hcup-us.ahrq.gov/db/nation/nis/nisdde.jsp

## Acknowledgement

The authors are extremely thankful to HCUP and AHRQ for allowing access to the database used in this study. As per their DUA, users who had access to the database underwent the appropriate training, and signed the required DUA. We are also thankful to HCUP’s partners and collaborators and the list can be found at https://www.hcup-us.ahrq.gov/partners.jsp.

## References

1. Ramphul K, Ramphul Y, Park Y, Lohana P, Dhillon BK, Sombans S. A comprehensive review and update on severe acute respiratory syndrome coronavirus 2 (SARS-CoV-2) and Coronavirus disease 2019 (COVID-19): what do we know now in 2021? Archives of medical sciences Atherosclerotic diseases. 2021;6:e5–e13.

2. Ramphul K, Mejias SG. Coronavirus Disease: A Review of a New Threat to Public Health. Cureus. 2020;12(3):e7276.

3. Holshue ML, DeBolt C, Lindquist S, Lofy KH, Wiesman J, Bruce H, et al. First Case of 2019 Novel Coronavirus in the United States. The New England journal of medicine. 2020;382(10):929–36.

4. Biancolella M, Colona VL, Mehrian-Shai R, Watt JL, Luzzatto L, Novelli G, et al. COVID-19 2022 update: transition of the pandemic to the endemic phase. Human genomics. 2022;16(1):19.

5. Bhopal SS, Bagaria J, Olabi B, Bhopal R. Children and young people remain at low risk of COVID-19 mortality. The Lancet Child & adolescent health. 2021;5(5):e12–e3.

6. HCUP National Inpatient Sample (NIS). Healthcare Cost and Utilization Project (HCUP). 2020. Agency for Healthcare Research and Quality, Rockville, MD. 2022 [Available from: http://www.hcup-us.ahrq.gov/nisoverview.jsp.

7. Kadri SS, Gundrum J, Warner S, Cao Z, Babiker A, Klompas M, et al. Uptake and Accuracy of the Diagnosis Code for COVID-19 Among US Hospitalizations. Jama. 2020;324(24):2553–4.

8. Parcha V, Booker KS, Kalra R, Kuranz S, Berra L, Arora G, et al. A retrospective cohort study of 12,306 pediatric COVID-19 patients in the United States. Scientific reports. 2021;11(1):10231.

9. COVID-19 Cases and Deaths by Race/Ethnicity: Current Data and Changes Over Time 2022 [Available from: https://www.kff.org/coronavirus-covid-19/issue-brief/covid-19-cases-and-deaths-by-race-ethnicity-current-data-and-changes-over-time/.

10. Goyal MK, Simpson JN, Boyle MD, Badolato GM, Delaney M, McCarter R, et al. Racial and/or Ethnic and Socioeconomic Disparities of SARS-CoV-2 Infection Among Children. Pediatrics. 2020;146(4).

11. Isath A, Malik AH, Goel A, Gupta R, Shrivastav R, Bandyopadhyay D. Nationwide Analysis of the Outcomes and Mortality of Hospitalized COVID-19 Patients. Current Problems in Cardiology. 2023;48(2):101440.

12. Rosenthal N, Cao Z, Gundrum J, Sianis J, Safo S. Risk Factors Associated With In-Hospital Mortality in a US National Sample of Patients With COVID-19. JAMA network open. 2020;3(12):e2029058.

13. Oliveira E, Parikh A, Lopez-Ruiz A, Carrilo M, Goldberg J, Cearras M, et al. ICU outcomes and survival in patients with severe COVID-19 in the largest health care system in central Florida. PloS one. 2021;16(3):e0249038.

14. Gonzalez-Dambrauskas S, Vasquez-Hoyos P, Camporesi A, Cantillano EM, Dallefeld S, Dominguez-Rojas J, et al. Paediatric critical COVID-19 and mortality in a multinational prospective cohort. Lancet regional health Americas. 2022;12:100272.

15. Penwill NY, Roessler De Angulo N, Pathak PR, Ja C, Elster MJ, Hochreiter D, et al. Changes in pediatric hospital care during the COVID-19 pandemic: a national qualitative study. BMC health services research. 2021;21(1):953.

16. Paquette ET, Derrington S, Fry JT, Michelson K, Patel A, Shah S, et al. Shifting Duties of Children’s Hospitals During the COVID-19 Pandemic. Journal of hospital medicine. 2020;15(10):631–3.

17. Tasnim S, Hossain MM, Mazumder H. Impact of Rumors and Misinformation on COVID-19 in Social Media. Journal of preventive medicine and public health = Yebang Uihakhoe chi. 2020;53(3):171–4.

18. Guidry JPD, Miller CA, Ksinan AJ, Rohan JM, Winter MA, Carlyle KE, et al. COVID-19-Related Misinformation among Parents of Patients with Pediatric Cancer. Emerging infectious diseases. 2021;27(2):650–2.

19. Aleem A, Akbar Samad AB, Slenker AK. Emerging Variants of SARS-CoV-2 And Novel Therapeutics Against Coronavirus (COVID-19). StatPearls. Treasure Island (FL): StatPearls Publishing Copyright © 2022, StatPearls Publishing LLC.; 2022.

20. Kong G, Kuguru KE, Krishnan-Sarin S. Gender Differences in U.S. Adolescent E-Cigarette Use. Current addiction reports. 2017;4(4):422–30.

21. Dana R, Bannay A, Bourst P, Ziegler C, Losser MR, Gibot S, et al. Obesity and mortality in critically ill COVID-19 patients with respiratory failure. International journal of obesity (2005). 2021;45(9):2028–37.

22. Albashir AAD. The potential impacts of obesity on COVID-19. Clinical medicine (London, England). 2020;20(4):e109–e13.

23. Harwood R, Yan H, Talawila Da Camara N, Smith C, Ward J, Tudur-Smith C, et al. Which children and young people are at higher risk of severe disease and death after hospitalisation with SARS-CoV-2 infection in children and young people: A systematic review and individual patient meta-analysis. EClinicalMedicine. 2022;44:101287.

24. Preston LE, Chevinsky JR, Kompaniyets L, Lavery AM, Kimball A, Boehmer TK, et al. Characteristics and Disease Severity of US Children and Adolescents Diagnosed With COVID-19. JAMA network open. 2021;4(4):e215298.

25. Smitherman LC, Golden WC, Walton JR. Health Disparities and Their Effects on Children and Their Caregivers During the Coronavirus Disease 2019 Pandemic. Pediatric clinics of North America. 2021;68(5):1133–45.

26. Rakus A, Soni A. Association between state Medicaid expansion status and health outcomes during the COVID-19 pandemic. Health services research. 2022;57(6):1332–41.

27. Mena GE, Martinez PP, Mahmud AS, Marquet PA, Buckee CO, Santillana M. Socioeconomic status determines COVID-19 incidence and related mortality in Santiago, Chile. Science (New York, NY). 2021;372(6545).

28. Li SL, Pereira RHM, Prete CA, Jr., Zarebski AE, Emanuel L, Alves PJH, et al. Higher risk of death from COVID-19 in low-income and non-White populations of São Paulo, Brazil. BMJ global health. 2021;6(4).

29. Abedi V, Olulana O, Avula V, Chaudhary D, Khan A, Shahjouei S, et al. Racial, Economic, and Health Inequality and COVID-19 Infection in the United States. Journal of racial and ethnic health disparities. 2021;8(3):732–42.

